# HaptiKart: An engaging videogame reveals elevated proprioceptive bias in individuals with autism spectrum disorder

**DOI:** 10.1101/2025.01.10.25320221

**Authors:** Daniel E. Lidstone, Mohit Singhala, Liam J. Wang, Jeremy D. Brown, Stewart H. Mostofsky

**Affiliations:** School of Behavioral Sciences and Education, Penn State Harrisburg, Middletown, Pennsylvania, United States of America; Center for Neurodevelopmental and Imaging Research, Kennedy Krieger Institute, Baltimore, Maryland, United States of America; Department of Otolaryngology-Head & Neck Surgery, University of Pittsburgh, Pittsburgh, Pennsylvania, United States of America; Electrical Engineering and Computer Science Department, University of Michigan, Ann Arbor, Michigan, United States of America; Department of Mechanical Engineering, Johns Hopkins University, Baltimore, Maryland, United States of America; Department of Neurology, Johns Hopkins University School of Medicine, Baltimore, Maryland, United States of America; Department of Psychiatry and Behavioral Sciences, Johns Hopkins University School of Medicine, Baltimore, Maryland, United States of America

## Abstract

An overreliance on proprioceptive (intrinsic) sensory input from the body, compared to visual (extrinsic) input from the environment, may underpin core features of autism spectrum disorder (ASD). We developed an engaging videogame (“HaptiKart”) as a tool to examine differences in sensory-motor bias (proprioceptive vs. visual) in children and adults with ASD and whether bias correlates with age, core autism features, and intellectual ability. Eighty-one participants (33 ASD, 48 typically-developing, TD) aged 8 to 31 years played “HaptiKart,” a driving videogame with a force-feedback steering wheel that provided “steering assist” during gameplay. In separate trials, proprioceptive and visual feedback were selectively delayed, and differences in driving error between the conditions were used to calculate perceptual bias scores. Effects of autism diagnosis and age on bias scores were examined, controlling for sex, as were associations of perceptual bias with autism symptom severity (ADOS-2, SRS-2), attention-deficit symptom severity (Conners4 ADHD Total Scores) ratings, and IQ (general ability index, GAI). The ASD group exhibited significantly higher proprioceptive bias than did the TD group (p=0.002). There was a trend for decreasing proprioceptive bias with age, but no significant diagnosis-by-age interaction. Increased proprioceptive bias correlated with higher autism severity and with lower IQ, but not ADHD symptoms. HaptiKart provides a highly scalable approach for measuring sensory-motor bias, revealing that individuals with ASD show elevated proprioceptive bias, correlating with autism severity. HaptiKart’s sensory-motor bias measure may thereby serve as a digital biomarker for addressing autism heterogeneity in ways that can improve targeted intervention.

**Author Summary:** In this study, we explored how people with autism spectrum disorder (ASD) may rely more on body-based (proprioceptive) senses than on visual information when interacting with their surroundings. Using a custom video game, *HaptiKart*, we designed a fun and engaging driving task to measure this sensory preference. Players drove a virtual car using a steering wheel that could be set to delay either visual or proprioceptive feedback, allowing us to observe how each type of feedback affects driving accuracy.

Our results showed that individuals with ASD had a stronger preference for proprioceptive feedback than those without ASD. This sensory preference was linked to greater autism symptom severity and lower IQ, suggesting that heightened reliance on body-centered feedback may contribute to learning and skill differences often seen in autism. These findings support the potential of *HaptiKart* as a simple, accessible tool to help clinicians better understand and address sensory biases in ASD, tailoring interventions to improve learning from visual information.

## Introduction

Acquiring social-communicative and motor skills from others through observation and imitation requires efficient dynamic visual-motor integration (VMI). Evidence from children with autism spectrum disorder (ASD) shows significant differences in dynamic VMI in non-social (e.g., ball catching) and social (e.g., motor imitation) behaviors, which may affect the implicit learning of motor and social skills [1,2]. Studies suggest a shared neural basis underlying both motor and social-communicative differences in children with ASD, demonstrated by differences in tasks that rely on dynamic VMI, such as ball catching, motor imitation, and visual-motor tracking [3–6]. Moreover, differences in dynamic VMI can be detected as early as six months of age, potentially serving as an early indicator of ASD [7]. This suggests that differences in brain mechanisms responsible for dynamic VMI may be present at birth or at least emerge early in children who later develop the core autism phenotype and receive an ASD diagnosis.

Autism-associated challenges with dynamic visual-motor integration (VMI) may lead to a preference for intrinsic proprioceptive feedback over extrinsic visual feedback in daily tasks. This bias could support skill acquisition in static environments but impede learning through observation in dynamic settings. Research shows that dynamic visual input perturbations disrupt motor control less in children with ASD compared to typically developing peers, indicating a stronger reliance on proprioceptive input [8–10]. Importantly, this elevated proprioceptive bias seems unique to ASD, as it is not seen in conditions like ADHD [11]. While elevated proprioceptive bias is not a core diagnostic features of ASD, this anomalous sensory-motor bias may serve as an important early indicator of ASD, useful for both diagnosis and guiding precision interventions. Findings also suggest that while elevated proprioceptive bias is observed in younger children with ASD, under 12 years, these biases may diminish in adolescence [9,10,12], potentially influenced by participation in interventions that promote learning from dynamic visual cues, such as Social Skills Groups.

In addition to disrupted social skills, a prolonged period of elevated proprioceptive bias during childhood may also affect intellectual development by disrupting implicit learning from visual stimuli in the environment. Individuals with lower intellectual abilities may therefore rely more on proprioceptive feedback for environmental exploration, potentially limiting their learning experiences. However, the literature has yet to explore whether elevated proprioceptive bias correlates with intellectual differences in individuals with ASD..

The relative weighting of proprioceptive versus visual feedback is a simple measure that may serve as a useful digital biomarker of a particular autism subtype with utility for aiding diagnosis, parsing autism heterogeneity, and guiding intervention. However, previous computational methods to examine proprioceptive bias have relied on inaccessible tools (i.e., expensive, non-portable robots) and long-duration lab assessments that limit clinical utility and accessibility [11,13,14]. Furthermore, proprioceptive bias has primarily been examined using tasks, such as ballistic reaching, where proprioception is the primary feedback modality [10,11,13–15]. Thus, it remains unclear whether sensory-motor bias is also present in visual-dominant tasks in individuals with ASD.

To address limitations in prior computational methods for examining sensory-motor bias, we developed “HaptiKart,” an arcade-style driving videogame (**Fig 1**). While existing videogame-like applications have been developed specifically for children with ASD using off-the-shelf technologies [16,17], there is still a gap in the availability of tools that specifically measure sensory-motor biases. Current methods largely focus on cognitive and behavioral aspects, often neglecting the role of sensory processing in ASD, particularly how reliant individuals with ASD are on proprioceptive feedback as compared to visual feedback during motor tasks. This study aims to address that gap by introducing a novel approach for assessing sensory-motor bias through a customizable and engaging videogame, HaptiKart. Unlike traditional clinical tools, which can be time-consuming and limited to controlled environments, HaptiKart provides a scalable solution that is both accessible and engaging for individuals with ASD.

**Fig 1.**
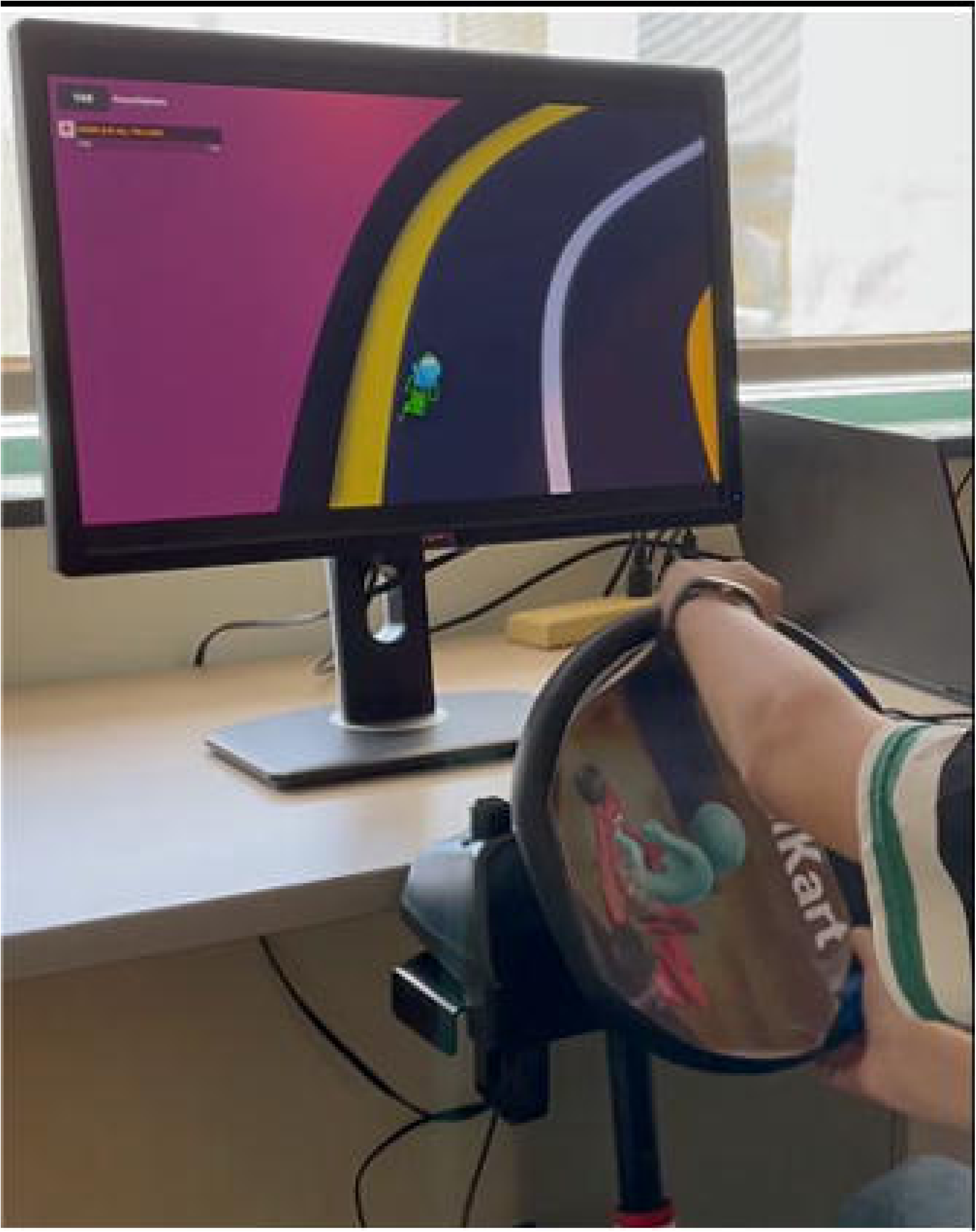
HaptiKart videogame. A child participant with ASD diagnosis playing the HaptiKart videogame.

The HaptiKart videogame assesses sensory-motor bias by delivering configurable visual feedback on the car’s position and proprioceptive feedback through a force-feedback steering wheel and selectively disrupting each feedback modality in two conditions—delayed proprioceptive feedback and delayed visual feedback. Importantly, this approach to delivering proprioceptive and visual error was implicit, as it was challenging for participants to recognize whether the visual or proprioceptive feedback had changed explicitly. Additionally, the order of the conditions was counterbalanced within each group to control for potential order effects.

This first study of HaptiKart had three aims: (1) to examine the effects of ASD diagnosis on perceptual sensory-motor (proprioceptive vs. visual) bias, (2) to examine the effects of age, as well as age-by-diagnosis on perceptual bias, (3) to examine associations with ASD clinical severity, as well as with severity of commonly co-occurring ADHD symptoms and with intellectual ability. Our corresponding hypotheses were: (1) individuals with ASD would show elevated proprioceptive bias relative to TD individuals, (2) elevated proprioceptive bias would show a significant decrease with age with steeper age-related declines in the ASD vs. TD groups, and (3) elevated proprioceptive bias would correlate significantly with core autism features, but not attentional (ADHD) traits. We also hypothesized that, in addition to the association between elevated proprioceptive bias and core autism features, this bias would also be linked to lower intellectual ability. Specifically, we propose that disrupted visual input from the external environment, beyond just social stimuli, may more generally affect cognitive development.

## Results

### Elevated Proprioceptive Bias in Individuals with ASD Relative to TD Controls

To quantify sensory-motor bias, we calculated the bias score as the difference in driving errors between the counterbalanced delayed proprioceptive feedback condition (Error*_V+DP_*) and the delayed visual feedback condition (Error*_DV+P_*) (*Bias score =* Error*_V+DP_ -* Error*_DV+P_*). A positive bias score indicates a preference for proprioceptive feedback, while a negative score reflects a greater reliance on visual feedback (**Fig 2**).

**Fig 2.**
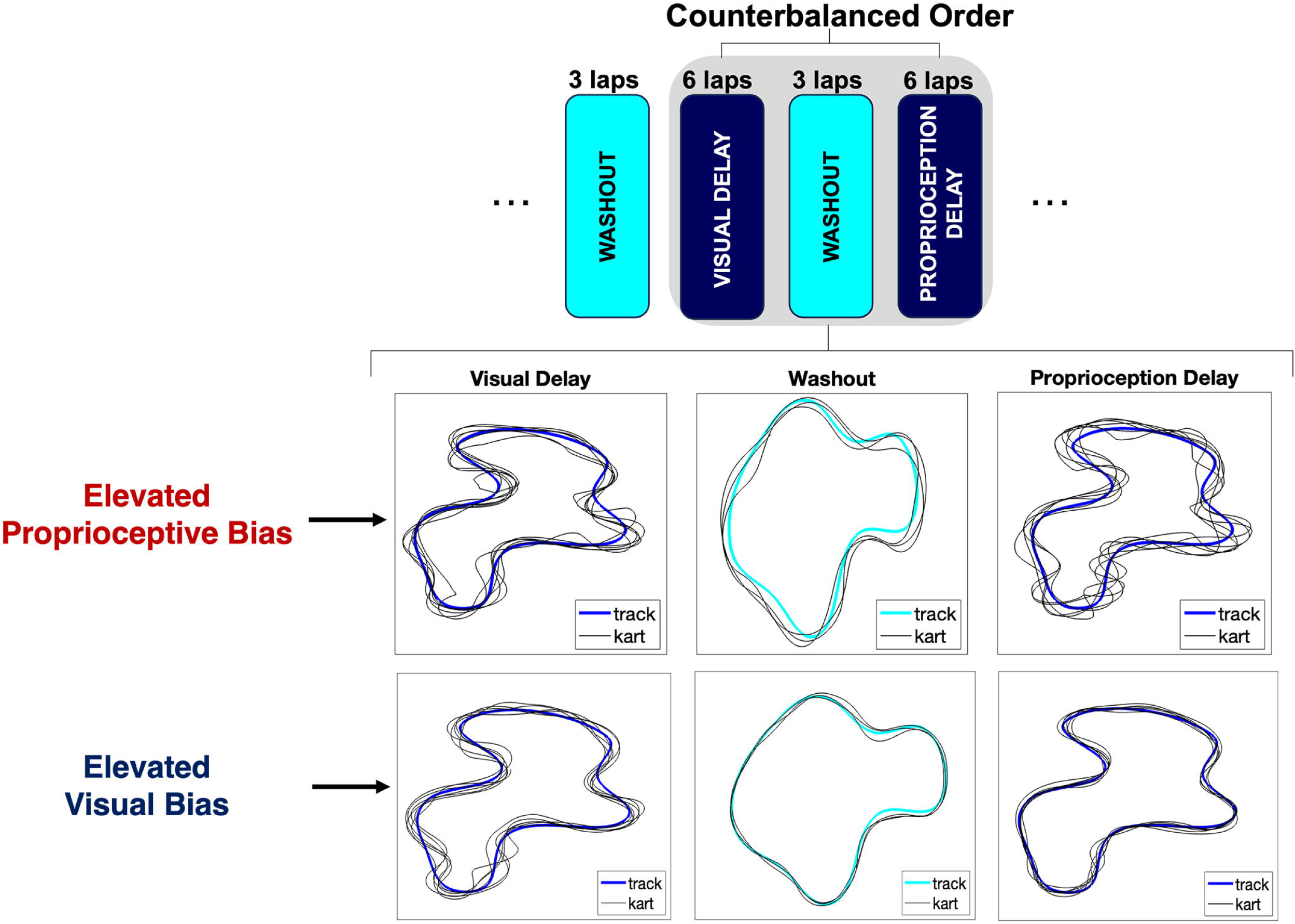
Experimental paradigm and sample data. Schematic showing the paradigm used to examine sensory-motor bias. The delayed visual and proprioceptive feedback conditions were counterbalanced within each group. Trials from two sample participants showing elevated proprioceptive bias (bias score = 7.85) and visual bias (bias score = -10.74).

The findings from ANCOVA did not show a significant diagnosis-by-age effect (p = 0.68) or effect of age (p = 0.06) but did show a significant effect of diagnosis (p = 0.002) on bias scores (**Fig 3**).

**Fig 3.**
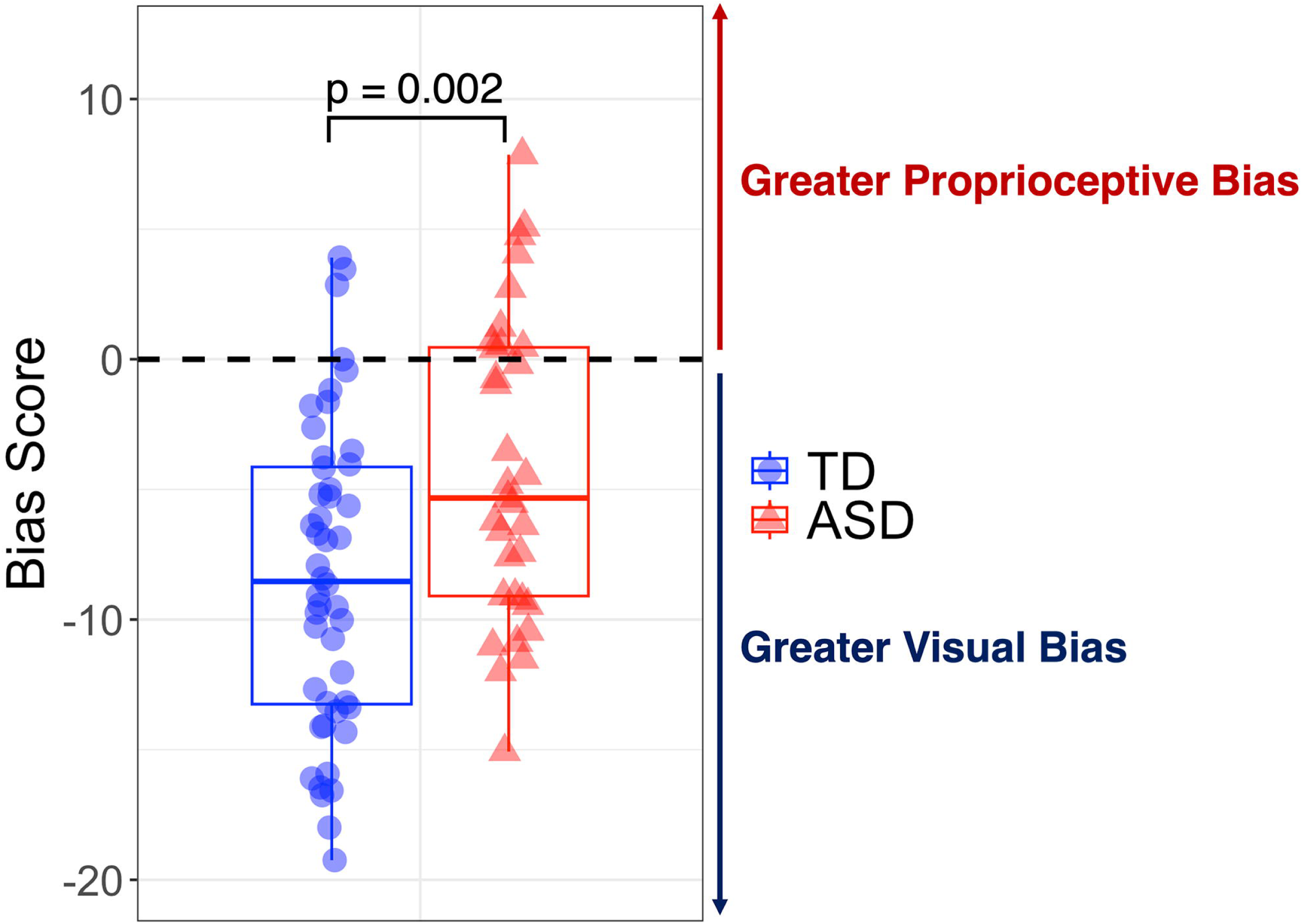
Group differences in perceptual bias scores. Children with ASD show significantly greater proprioceptive bias as compared to TD children.

Bias scores in the ASD group were more positive than the TD group (ASD bias score: -4.24 ± 5.83 vs. TD bias score: -8.34 ± 5.90), showing elevated proprioceptive bias. Further, the biological sex covariate did not have a significant effect on bias scores (p = 0.68). Collectively, individuals with an ASD diagnosis showed increased preference for proprioceptive feedback that was not explained by age or biological sex.

### Sensory-Motor Bias Correlates with Core Autism Features and IQ, but not ADHD Features

Correlations between sensory-motor bias and core autism features revealed significant correlation with clinical ratings of autism symptoms within the ASD group (ADOS-CS, r = 0.48, p = 0.035, p_adj_ = 0.046) and parent rated autism symptoms at the whole group level (SRS-2 total score, r = 0.52, p = 0.0003, p_adj_ = 0.0006) (**Fig 4A-B**), such that elevated symptoms predicted elevated proprioceptive bias. In contrast, parent-rated ADHD symptom severity was not significantly correlated with bias scores (Conners4 total score, r = 0.17, p = 0.38, p_adj_ = 0.38) (**Fig 4F**). Bias scores were also significantly correlated with GAI, such that elevated proprioceptive bias was associated with lower GAI (r = 0.54, p = 0.000002, p_adj_ = 0.000008) (**Fig 4C**).

**Fig 4.**
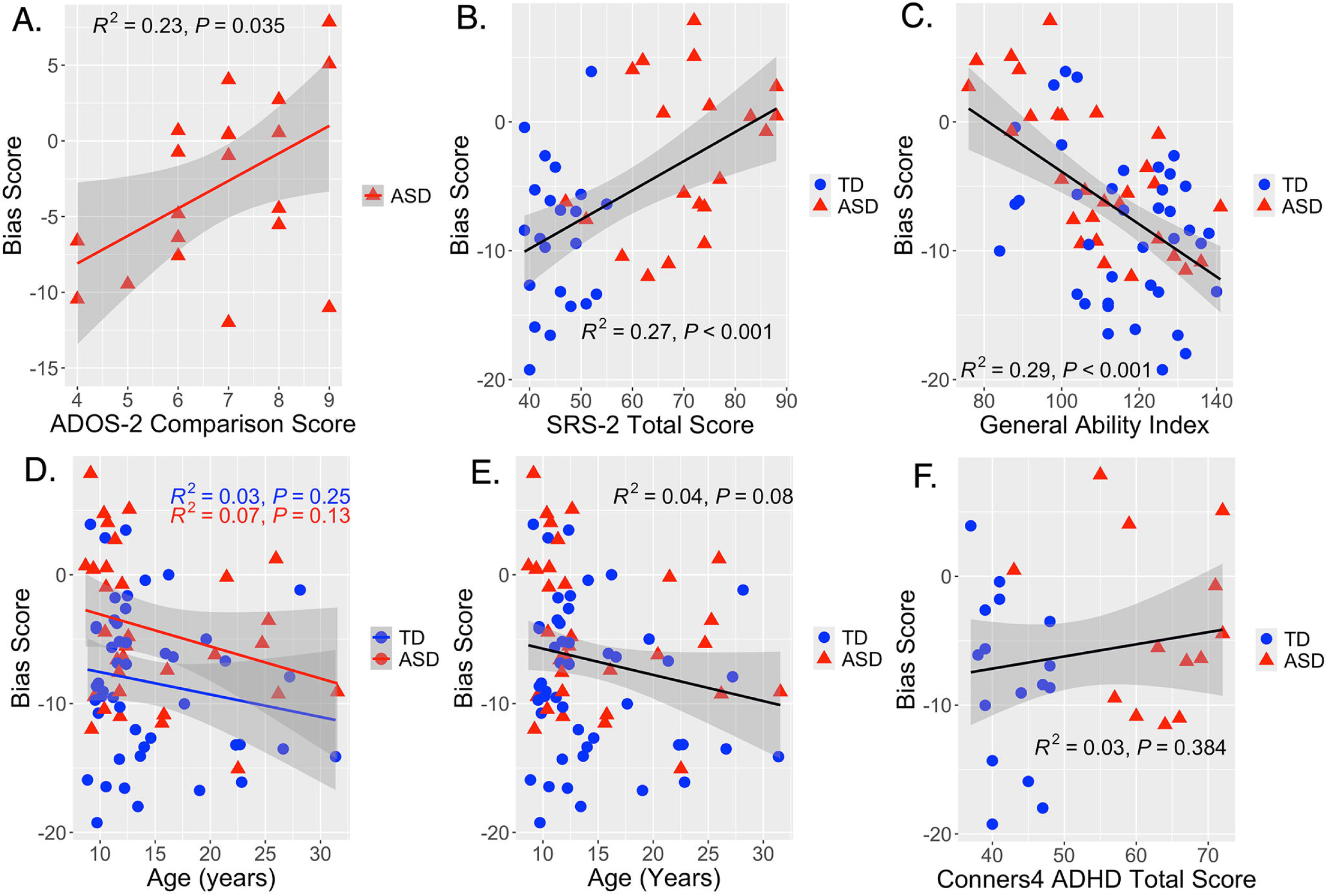
Correlations between perceptual bias scores, age, and autism-associated features: Simple linear regression showing a significant correlation between elevated proprioceptive bias and: (A) elevated clinically rated ASD symptoms (ADOS-CS (Module 3) scores, p=0.03), (B) elevated parent-rated ASD symptoms (SRS-2 total scores, p < 0.001), and (C) lower intellectual ability (GAI, p < 0.001). (D-E) Correlations between age and bias scores were not significant for either group (p>0.1) but showed a negative trend-level association at the whole cohort level (age, p=0.08). (F) There was no correlation between parent-rated ADHD symptoms and bias scores (Conners4 total score, p=0.38).

### Trend for Decreasing Proprioceptive Bias with Increasing Age Across Both ASD and TD Cohorts

ANCOVA revealed a trend-level main effect of age on bias scores (p = 0.06). Simple linear regression (bias scores ∼ age) revealed a trend for an effect of age on perceptual bias across the entire cohort, with decreasing proprioceptive bias with increasing age (r = -0.2, p = 0.08) (**Fig 4E**). However, there was no significant interaction of diagnosis and age (p = 0.68), such that this trend for increasing reliance on visual error vs. proprioceptive error from childhood to adulthood was similar in both diagnostic groups (ASD: r = -0.26, p=0.13; TD: r = -0.17, p = 0.25) (**Fig 4D**).

## Discussion

This study reveals two main findings that support the stated hypotheses: (1) children with ASD show significantly elevated proprioceptive bias relative to TD children, (2) elevated proprioceptive bias correlates with greater core autism clinical severity and with lower intellectual ability but not with ADHD clinical severity. While associations between age and proprioceptive bias were not significant, trend-level associations with age were observed such that elevated proprioceptive bias was observed in younger vs. older participants. Examination of diagnosis-by-age effects on sensory-motor bias did not support our hypothesis of elevated proprioceptive bias in younger but not older individuals with ASD vs. TD. These findings suggest that ASD-associated elevated proprioceptive bias persists across development and does not appear to normalize to TD levels with increasing age. However, including younger participants (<8 years of age) in future studies will provide a deeper insight into autism-associated differences in developmental trajectories of sensory-motor bias.

The finding that children with ASD show elevated proprioceptive bias in the current study supports a degree of task invariance of the measure as it has been previously observed in proprioception-dominant tasks such as ballistic reaching [11,13,14] and now with HaptiKart, a more visual task. Consistent with prior studies, the current study showed elevated proprioceptive bias correlated with parent-rated core autism features [13]. The finding that elevated proprioceptive bias correlated with core autism symptoms but not ADHD symptoms suggests that proprioceptive bias may be a promising digital phenotypic marker of autism.

Our findings indicate a strong correlation between preference for proprioceptive vs. visual feedback and general intellectual ability (GAI), suggesting this bias may be more pronounced in individuals with severe ASD symptoms. While previous studies have linked intellectual ability and motor skills in ASD, these correlations are expected due to the complexity of standardized motor tests, which can be challenging for those with autism and intellectual differences [18,19]. Notably, our study highlights a robust association between sensory-motor bias and IQ, focusing solely on sensory preferences rather than motor skills. This sensory-motor preference could partially explain intellectual differences in individuals with ASD and may serve as a target for therapies aimed at improving skill learning through visual observation and imitation. The HaptiKart task, requiring minimal instructions (“keep the car on the track centerline”) and being child-friendly, shows promise for reliably assessing sensory-motor preferences in the diverse ASD population. The strong link between greater proprioceptive bias and lower IQ also suggests that intervention approaches that aim to increase reliance on dynamic visual feedback and stimuli, particularly in early development, may enhance cognitive development.

HaptiKart’s ability to assess sensory-motor preferences makes it a valuable asset for clinicians and therapists. By identifying elevated proprioceptive bias, HaptiKart can help tailor interventions that focus on increasing reliance on visual feedback. This approach aligns with evidence-based autism therapies where therapists help guide children towards dynamic visual stimuli to teach social skills (e.g., eye/facial/limb movements) and language skills (e.g., mouth movement).

Therapists can also use HaptiKart’s quick assessments of sensory bias to develop therapy plans to modify sensory bias and as an objective outcome measure of treatment response following intervention. For children showing elevated proprioceptive bias, digital interventions can be designed to enhance visual feedback reliance by introducing inaccuracies in proprioceptive feedback, such as gradually reducing the gain of proprioceptive feedback. By fine-tuning visual and proprioceptive feedback, therapists can create diverse therapy goals tailored to individual needs. HaptiKart could be used to deliver interventions aimed at reducing anomalous sensory-motor bias at-home, reducing costs, and increasing access of services to families living in underserved and rural communities. Further, HaptiKart offers an objective measure of sensory bias, enabling therapists to monitor progress and adjust treatments as needed. Its digital format also ensures that these interventions are accessible across different cultures, unaffected by language barriers.

HaptiKart’s quick measure of proprioceptive bias can also provide clinicians and therapists information on how best to instruct a child on a new skill. For instance, children with elevated proprioceptive bias may benefit from a “hands-on” manual guidance approach when learning new skills (e.g., a therapist manually guides a child’s arm towards the target) as compared to using visual demonstration of the desired action. Therefore, HaptiKart has strong potential as an assessment and digital intervention tool that can be easily integrated within clinical settings to aid development of intervention plans to address anomalous sensory-motor biases.

While allowing for more targeted therapeutic intervention, future iterations of HaptiKart’s assessment of proprioceptive/tactile vs. visual bias may be extended to examine other types of sensory-motor biases such as proprioceptive vs. auditory and tactile vs. visual that can inform clinical practice. Further, an adapted version of HaptiKart could be extended to toddlers (1-3 years) to better understand typical developmental trajectories of sensory-motor bias with the goal of improving early identification and support for affected children during the most sensitive developmental period.

One limitation of our study is the lack of data on participants’ prior experiences with intervention services, which may influence sensory preferences. Future research should explore how these experiences relate to proprioceptive and visual biases. Additionally, we did not include a clinical control group (e.g., children with attention-deficit hyperactivity disorder), limiting our ability to assess the specificity of these biases to ASD. The single-session design also prevented assessment of the test-retest reliability of HaptiKart’s measures, which is important for clinical utility. Some participants were excluded for hand removal from the wheel, and future versions will incorporate sensors to capture this behavior. While using an off-the-shelf steering wheel enhances HaptiKart’s scalability, it lacks precise research-grade force feedback and has restricted applicability to broader and more complex motor tasks (e.g., reaching in 3D space). Future work will explore the use of research-grade haptic devices with two or three degrees of freedom to assess more complex motor behaviors. Another limitation is the lack of direct measurements of visual attention, which could affect sensory-motor bias. Future studies could incorporate eye-tracking or other methods to gain a better understanding of the relationship between visual attention and sensory-motor bias in ASD. Finally, the fixed visual (250 ms) and proprioceptive (300 ms) delays utilized in this study were grounded in prior research and pilot testing. Future studies could explore the use of dynamic or varied delays as they might offer additional insights into how sensory integration adjusts to varying temporal conditions and affects sensory bias.

In summary, our study using HaptiKart demonstrated significantly elevated proprioceptive bias in children with ASD compared to typically developing controls. The brief assessment format (approximately 5 minutes) has strong potential for clinical and home applications to track sensory preferences throughout development and guide interventions aimed at enhancing preference for dynamic visual feedback while reducing reliance on proprioceptive feedback in individuals with ASD.

## Materials and Methods

### Participants

A total of 81 participants 8-31 years old (33 ASD and 48 TD) participated in the study and met criteria for inclusion. Groups did not differ in age (ASD: 14.66 ± 6.28 years; TD: 14.49 ± 5.64 years; p = 0.9) or sex (ASD: 69% male, TD: 50% male; χ^2^=3.11, p = 0.07) (**Table 1**). Not included in these participant totals is one outlier in the ASD group with a bias score exceeding 3 standard deviations (SD) from the ASD group mean. An additional ASD participant was removed due to an outlier IQ score approximating 3 SD below the ASD mean. Additional participants not included in the study were eight participants (6 ASD and 2 TD children) that removed their hands from the wheel during gameplay. These participants had to be excluded as there was no objective way to determine at what interval their hands were removed from the wheel during gameplay.

**Table 1:** Participant demographics for all participants included in analysis.

|  | TD<br>(*n* = 48) | ASD<br>(*n* = 33) | Test Statistic |
| --- | --- | --- | --- |
| Age (years) | 14.49 ± 5.64 | 14.66 ± 6.28 | *p* = 0.90 |
| Sex (% male) | 50% | 70% | $\chi^2=3.11$, *p* = 0.07 |
| GAI<br>(TD: 38, ASD: 29) | 116.11 ± 15.08 | 108.66 ± 16.85 | *p* = 0.06 |
| SRS-2 Total Score<br>(TD: 22, ASD: 20) | 45.45 ± 4.81 | 70.30 ± 11.39 | *p* < 0.0001 |
| Conners4 Total Score<br>(TD: 16, ASD: 13) | 42.56 ± 4.03 | 62.92 ± 8.23 | *p* < 0.0001 |
| ADOS-2 CS (Module 3)<br>(ASD: 20) | - | 6.85 ± 1.50 | - |
| ADOS-2 Total Score<br>(Modules 3 and 4)<br>(ASD: 26) | - | 11.15 ± 3.07 | - |

TD: Typically developing
ASD: Autism spectrum disorder
GAI: General Ability Index
SRS-2: Social Responsiveness Scale, 2<sup>nd</sup> Edition
ADOS-2 CS: Autism Diagnostic Observation Schedule, 2<sup>nd</sup> Edition Comparison Score

All participants in the ASD group received a clinical diagnosis of ASD based on DSM-5 criteria, as confirmed by a pediatric neurologist with decades of experience working with individuals with ASD across both clinical and research settings (SHM). Child participants with ASD (8 to 17 years; n=25) were required to have a prior diagnosis of ASD, score within the autism spectrum range on the Autism Diagnostic Observation Schedule, 2nd edition (ADOS-2), and score within the clinical range on the Social Responsiveness Scale, 2^nd^ edition (SRS-2). Adult participants (>17 years; n=8) were required to have a prior clinical diagnosis of autism or ASD and score within the clinical range on the SRS-2. Further, the ADOS-2 Comparison Score (ADOS-CS) and SRS-2 were combined to analyze the clinical severity of ASD.

Additional data collected on subsets of participants included IQ General Ability Index (GAI) measured using the Wechsler Intelligence Scale for Children 5^th^ Edition (WISC-V) (participants < 12 years old) or Wechsler Abbreviated Scale of Intelligence 2^nd^ Edition (WASI-II) (participants >13 years old). For participants receiving the WISC-V, GAI was calculated using the Verbal Comprehension Index and Fluid Reasoning Index and converted to a composite score. For the participants completing the WASI-II, the full-scale IQ score calculated from Verbal Comprehension and Perceptual Reasoning subscales was used to calculate GAI. ADHD severity was assessed using the parent version of the Conners 4^th^ edition (Conners4) total score, derived from the inattention and hyperactivity/impulsivity subscales of the Conners4 and standardized using T-scores [20].

Ethics approval was received from the Johns Hopkins University School of Medicine Institutional Review Board before the study (IRB00269589). Written informed consent was obtained from all adult participants, legal guardians of each child participant, and verbal assent from all children.

### Experimental Design

#### HaptiKart Procedures

Participants played the custom-built HaptiKart videogame in a quiet, light-controlled room using a Logitech G29 steering wheel. The game was run on an MSI GS66 Stealth laptop and displayed on a Dell monitor adjusted to eye level. The wheel was affixed to a height-adjustable stand, with participants instructed to maintain their hands at a standardized 10 o’clock (left hand) and 2 o’clock (right hand) position (**Fig 1**). They were directed to “drive the car on the centerline” without awareness of any visual or steering perturbations. To ensure data reliability, participants who exhibited challenging behavior—specifically, removing their hands from the wheel during gameplay—were excluded from the analysis, as there was no objective measure for when hand release occurred. Six children with ASD and two TD children displayed this behavior and were not included in the analyses of participant characteristics or sensory preferences.

#### Measurement of Driving Error

During gameplay, errors were measured continuously as the mean of two error types: (1) the absolute distance of the car relative to the track centerline (position error, PE) that was expressed as a percentage of maximum lateral error (6 meters) and (2) the absolute orientation of the car relative to the track centerline (heading error, HE) that was expressed a percentage of the maximum orientation error (+/- 90-degrees) (**Fig 5**). The absolute relative position and heading errors were averaged across all laps within each block to produce a single error metric (Mean Total Error Percentage (%) = 0.5 x [ABS(PE)/6] + [ABS(HE)/90]).

**Fig 5.**
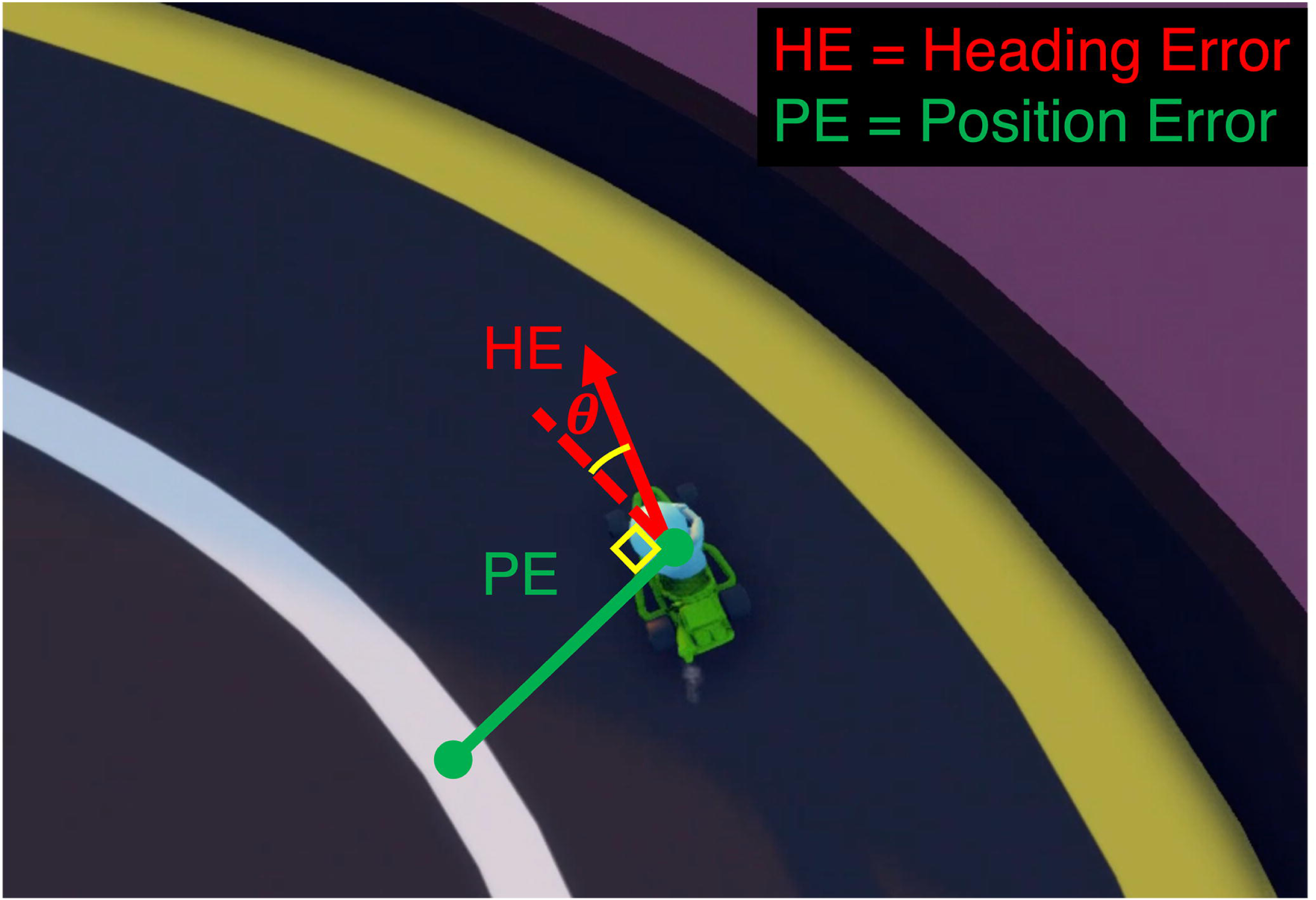
Error calculation. Continuous calculation of heading and position error used for proprioceptive feedback.

#### Visual-error feedback

The visual-error feedback (**Fig 6A**) was recognized as the car’s position relative to the track centerline. While other forms of error feedback could have been utilized—such as color changes or symbols indicating the direction and magnitude of error—the continuous feedback of the kart’s position relative to the centerline was most akin to the ongoing nature of proprioceptive feedback from the steering wheel. This similarity is crucial for quantifying sensory-motor bias because it allows for a direct comparison between the sensory modalities. By using a feedback mechanism that mimics the continuous flow of proprioceptive-error feedback, we can better assess how children with ASD integrate these two types of feedback. This approach ensures that any performance differences between the feedback delay conditions are more accurately attributed to differences in sensory bias rather than differences in the nature of the feedback provided.

**Fig 6.**
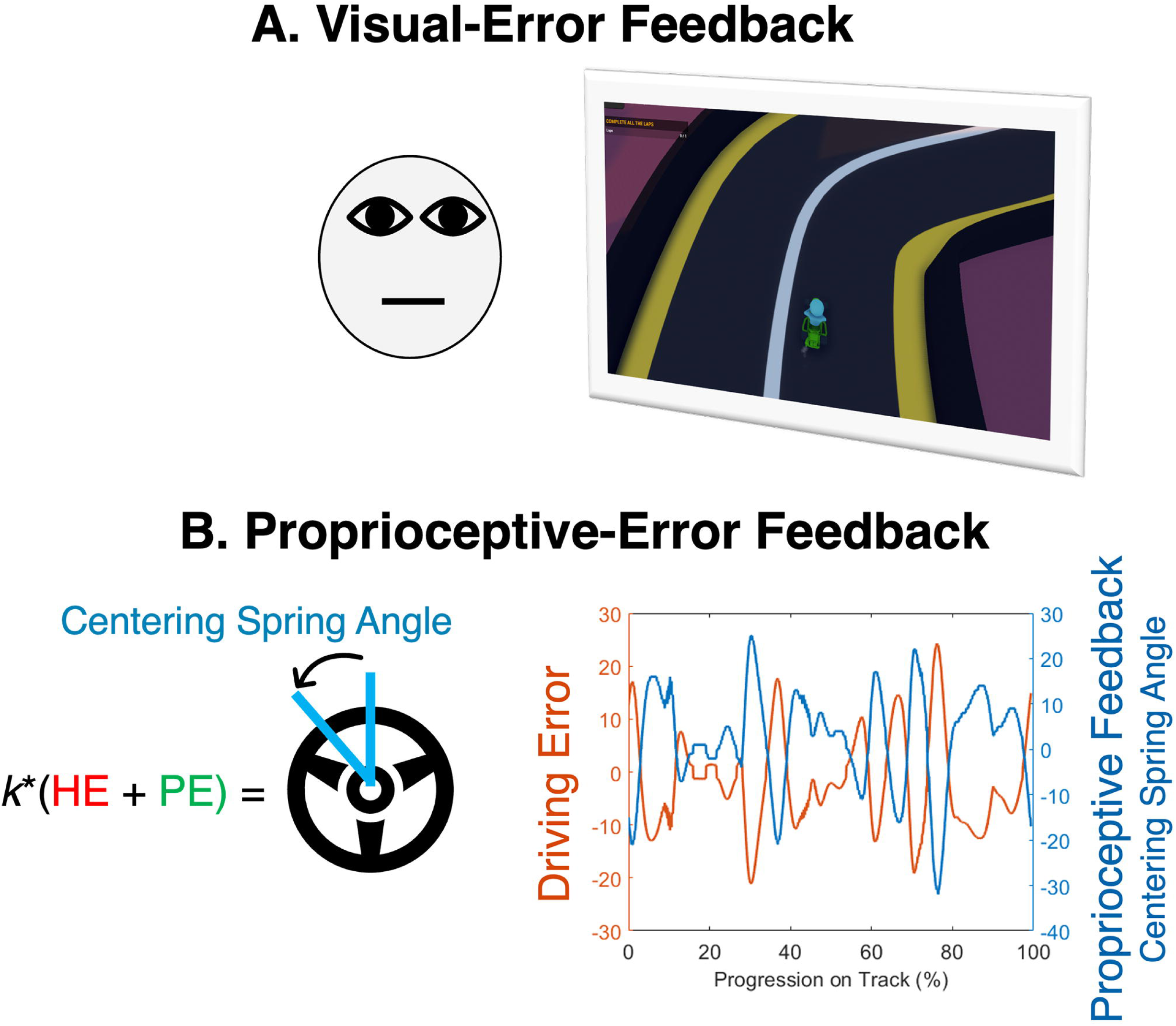
Visual and proprioceptive-error feedback. (A) Visual display seen by the participant during gameplay. (B) Method of delivering corrective proprioceptive-error feedback by using driving errors to update wheel centering spring angle (perceived as “steering assist”). Right plot in panel B shows opposing relationship between driving error and the corrective proprioceptive- error feedback.

#### Proprioceptive-error feedback (force feedback wheel)

The two error types (HE and PE) were used to provide proprioceptive-error feedback, felt as a corrective torque or “steering assist” applied to the wheel, that continuously corrected steering by guiding the car toward the track centerline. The gain constant (*k*) on the “steering assist” was multiplied by each error type (HE and PE) to adjust the resting angle of the centering spring of the steering wheel (**Eq. 1**, **Fig 6B**). The gain constant was selected to provide some “steering assistance” while still requiring steering input from the user to stay on the centerline (see supplement **Fig S3**). Detailed description of the proprioceptive-error feedback is provided in the supplement.

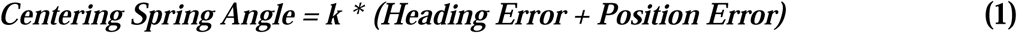

#### Temporally Delayed Feedback

The paradigm for examining sensory-motor bias involved selectively delaying proprioceptive and visual feedback in separate conditions, which allowed us to measure the relative preferences for each feedback modality. The chosen visual delay was 250 milliseconds (ms), while the proprioceptive delay was 300 ms. These delays were selected to ensure they could be reliably detected by young children [21]. Previous findings in children as young as five years old show that visual-proprioceptive asynchrony detection thresholds continue to increase up to 300 ms [21]. Pilot testing with a small group of children revealed that a 300 ms visual delay might be too challenging due to the visual dominance of the driving task, so a shorter 250 ms visual delay was selected.

#### Experimental Paradigm

The paradigm used to measure sensory-motor bias was completed as part of a larger paradigm that included several sensory conditions, completed in the same order for all participants, and separated by washout blocks (see supplement; **Fig S1**). The washout blocks provided veridical visual and proprioceptive feedback to mitigate crossover effects between conditions with varied sensory feedback. Below, we describe a “brief” paradigm (∼5-min) within the larger paradigm that was designed to examine sensory-motor bias while controlling for both condition order (i.e., counterbalancing) and crossover effects (i.e., using washout blocks).

Following several different sensory feedback conditions that all participants completed in a set order (**Fig S1**), each participant completed the brief paradigm to examine sensory-motor bias. The brief paradigm included three conditions: (1) visual delay (*DV+P*; 6 laps), (2) washout (3 laps), and (3) proprioceptive delay (*V+DP*; 6 laps) (6 laps) (**Fig 2**). The car’s velocity was fixed throughout gameplay (a top speed of 15.0 m/s and an acceleration of 10.0 m/s²), ensuring consistent track progression speed across participants, and the laps for each condition were completed consecutively (i.e., no rest between laps) with the condition ending once the car crossed the finish line at the conclusion of lap 6. The experimenter provided participants with ∼20-seconds of rest between conditions.

The average lap times (in seconds) were calculated for both the visual delay conditions (TD: 22.5 ± 1.85 sec; ASD: 23.1 ± 2.40 sec; *p* = 0.24) and proprioceptive delay conditions (TD: 20.8 ± 2.46 sec; ASD: 21.5 ± 2.06 sec; *p* = 0.18), with findings revealing no significant effects of diagnosis on completion times between groups for either delay condition (see supplemental **Fig S2**). While mean lap times did not significantly differ between the groups for either condition, variability in lap times was observed due to participant-specific responses during gameplay such as collisions with the guardrails along the track.

In addition to guardrails designed to prevent the vehicle from leaving the track, HaptiKart incorporates task constraints to maintain continuity and ensure progression along the track. A “wrong way” respawn system automatically resets the vehicle when driven in reverse, aligning it forward along the track centerline. Similarly, an “out of bounds” respawn mechanism was also implemented: if a sustained collision causes the car to leave the track, HaptiKart immediately resets its position and orientation, ensuring forward alignment on the centerline and preserving task continuity, similar to the “wrong way” reset.

More laps (6 vs. 3) were completed in the delay conditions to facilitate analyses of online learning during the delay conditions that could influence our primary outcome measure (i.e., sensory-motor bias). To ensure that errors were comparably stable between groups for each condition, differences in online learning were examined (Equation 2).

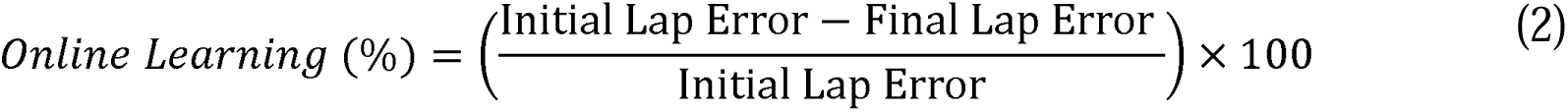

The findings revealed no significant differences in online learning between the ASD and TD groups (assessed as the decrease in error from the first to last lap) for either visual delay (TD: - 12.7 ± 35.0%; ASD: -6.45 ± 29.2%; *p* = 0.38) or proprioceptive delay (TD: 1.95 ± 30.2%; ASD: -4.66 ± 35.6%; *p* = 0.38) (see Supplement **Fig S4**). Since no differences were observed in learning for each delay condition between ASD and TD groups, the driving error data was aggregated across all six laps for each condition (visual delay: Error*_DV+P_*; proprioceptive delay: Error*_V+DP_*).

To account for potential order effects on the measurement of sensory-motor bias, the order of the delay conditions was counterbalanced within each group such that an equal proportion of participants completed the visual delay condition first and the other half completed the proprioceptive delay condition first. Due to the exclusion of some participants (see Participants section), the exact proportion of participants included in this study that completed the visual delay first was 46% and 45% for TD and ASD groups, respectively.

#### Calculation of Sensory-Motor Bias

Sensory-motor bias was calculated as the difference in mean total error between the delayed proprioceptive feedback (Error*_V+DP_*) and delayed visual feedback (Error*_DV+P_*) conditions, as shown in Equation (3), where a positive score (>0) indicates proprioceptive bias and a negative score (<0) indicates visual bias (**Fig 7**).

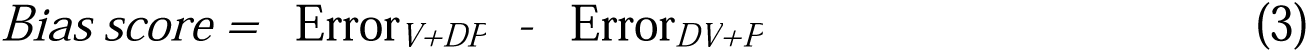

**Fig 7.**
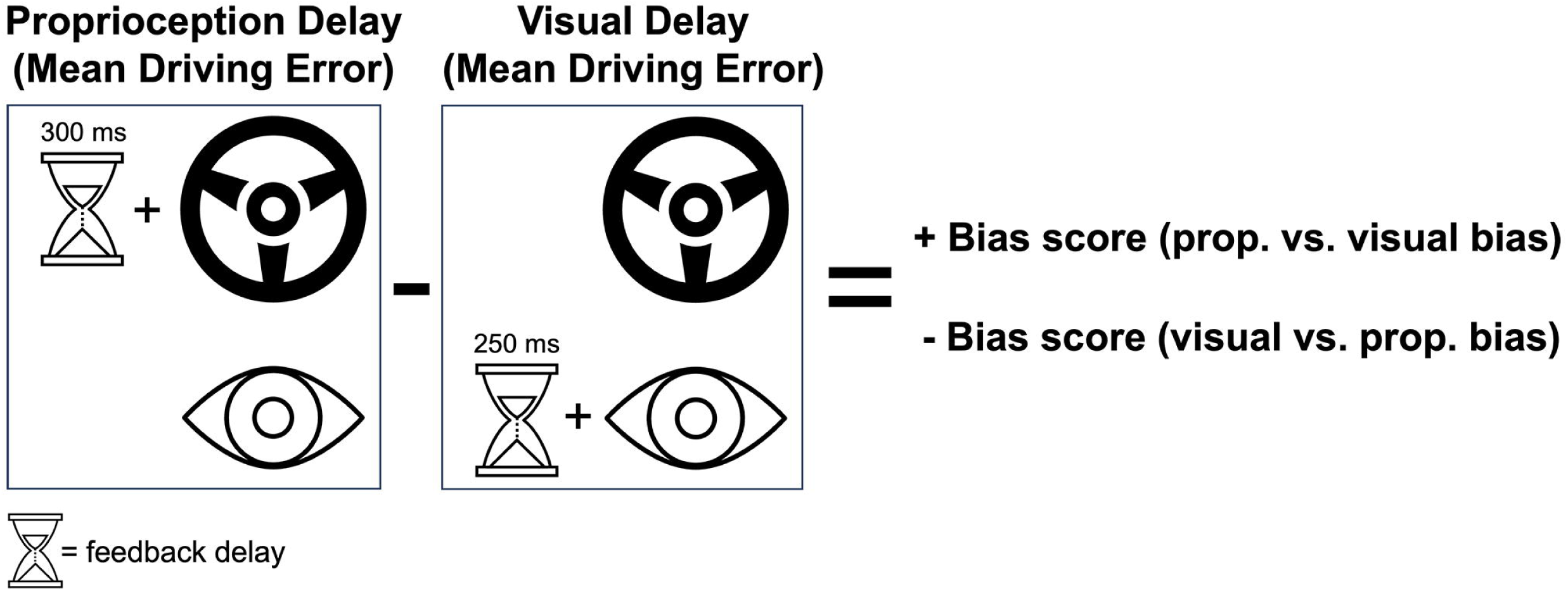
Calculation of perceptual bias scores. Visualization of the two delay conditions and interpretation of the bias score.

### Statistical analysis

To examine the interaction effects between bias scores and age, an ANCOVA was conducted to assess the interaction effect of diagnosis and age on bias scores while controlling for biological sex as a covariate (bias score ∼ diagnosis*age + sex). Overall main effects of diagnosis and age on bias scores were also evaluated. Simple linear regression was used to examine the correlation between bias scores and core autism symptoms (ADOS-CS and SRS-2 total score), ADHD symptoms (Conners4 total score), intellectual ability (GAI), and age. Significance for all tests was set at an alpha level of 0.05, and false discovery rate correction was applied to the p-values in the correlation analysis. All statistical analyses were conducted using RStudio version 2021.09.1. All assumptions for ANCOVA were met including homogeneity of variance (Levene’s Test: p = 0.7), and normality of residuals (Shapiro-Wilk; p = 0.10). Homogeneity of slopes was not assessed separately, as it is part of the hypothesis being tested.

## Data Availability

The data that support the findings of this study are available from the corresponding
author upon reasonable request.

## Acknowledgements

We sincerely thank our participants and their families for their time, effort, and support, which made this study possible.

## Supporting Information

**S1 File. Full experimental paradigm and supplemental figures.**

This document contains a detailed description of the full experimental paradigm used in the HaptiKart task, including block structure, feedback perturbations, steering wheel parameters, and track generation procedures. It also includes the following supplemental figures:

- **Figure S1.** Schematic of the experimental paradigm.
- **Figure S2.** Average lap times by group and condition.
- **Figure S3.** Driving paths under proprioceptive-error feedback without user input.
- **Figure S4.** Online learning effects by condition.
- **Figure S5.** Track generation using Bezier curves in Unity.
- **Figure S6.** Track configurations used in the full paradigm.

## References

1. Lidstone DE, Mostofsky SH. Moving Toward Understanding Autism: Visual-Motor Integration, Imitation, and Social Skill Development. Pediatr Neurol. 2021;122: 98–105. doi:10.1016/j.pediatrneurol.2021.06.010

2. Mostofsky SH, Ewen JB. Altered connectivity and action model formation in autism is autism. Neuroscientist. 2011;17: 437–448. doi:10.1177/1073858410392381

3. Ament K, Mejia A, Buhlman R, Erklin S, Caffo B, Mostofsky S, et al. Evidence for Specificity of Motor Impairments in Catching and Balance in Children with Autism. J Autism Dev Disord. 2015;45: 742–751. doi:10.1007/s10803-014-2229-0

4. De Francesco S, Morello L, Fioravanti M, Cassaro C, Grazioli S, Busti Ceccarelli S, et al. A multimodal approach can identify specific motor profiles in autism and attention-deficit/hyperactivity disorder. Autism Research. 2023. doi:10.1002/aur.2989

5. Lidstone DE, Miah FZ, Poston B, Beasley JF, Mostofsky SH, Dufek JS. Children with Autism Spectrum Disorder Show Impairments During Dynamic Versus Static Grip-force Tracking. Autism Research. 2020;13: 2177–2189. doi:10.1002/aur.2370

6. Tunçgenç B, Pacheco C, Rochowiak R, Nicholas R, Rengarajan S, Zou E, et al. Computerised Assessment of Motor Imitation (CAMI) as a scalable method for distinguishing children with autism. Biol Psychiatry Cogn Neurosci Neuroimaging. 2020;6: 321–328. doi:10.1016/j.bpsc.2020.09.001

7. Landa RJ, Haworth JL, Nebel MB. Ready, Set, Go! Low Anticipatory Response during a Dyadic Task in Infants at High Familial Risk for Autism. Front Psychol. 2016;7: 721. doi:10.3389/fpsyg.2016.00721

8. Gepner B, Mestre D, Masson G, de Schonen S. Postural effects of motion vision in young autistic children. Neuroreport. 1995;6: 1211–1214. doi:10.1097/00001756-199505300-00034

9. Greffou S, Bertone A, Hahler EM, Hanssens JM, Mottron L, Faubert J. Postural hypo- reactivity in autism is contingent on development and visual environment: A fully immersive virtual reality study. J Autism Dev Disord. 2012;42: 961–970. doi:10.1007/s10803-011-1326-6

10. Hirai M, Sakurada T, Izawa J, Ikeda T, Monden Y, Shimoizumi H, et al. Greater reliance on proprioceptive information during a reaching task with perspective manipulation among children with autism spectrum disorders. Sci Rep. 2021;11. doi:10.1038/s41598-021-95349-0

11. Izawa J, Pekny SE, Marko MK, Haswell CC, Shadmehr R, Mostofsky SH. Motor learning relies on integrated sensory inputs in ADHD, but over-selectively on proprioception in autism spectrum conditions. Autism Research. 2012;5: 124–136. doi:10.1002/aur.1222

12. Takamuku S, Ohta H, Kanai · Chieko, De AF, Hamilton C, Gomi H. Seeing motion of controlled object improves grip timing in adults with autism spectrum condition: evidence for use of inverse dynamics in motor control. Exp Brain Res. 2021;239: 1047–1059. doi:10.1007/s00221-021-06046-3

13. Haswell CC, Izawa J, Dowell LR, Mostofsky SH, Shadmehr R. Representation of internal models of action in the autistic brain. Nat Neurosci. 2009;12: 970–2. doi:10.1038/nn.2356

14. Marko MK, Crocetti D, Hulst T, Donchin O, Shadmehr R, Mostofsky SH. Behavioural and neural basis of anomalous motor learning in children with autism. Brain. 2015;138: 784–797. doi:10.1093/brain/awu394

15. Glazebrook CM, Gonzalez D, Hansen S, Elliott D. The role of vision for online control of manual aiming movements in persons with autism spectrum disorders. Autism. 2009;13: 411–433. doi:10.1177/1362361309105659

16. Wintner SR, Waters SE, Peechatka A, Gonzalez-Heydrich J, Kahn J. Evaluation of a scalable online videogame-based biofeedback program to improve emotion regulation: A descriptive study assessing parent perspectives. Internet Interv. 2022;28. doi:10.1016/j.invent.2022.100527

17. McCleery JP, Zitter A, Solórzano R, Turnacioglu S, Miller JS, Ravindran V, et al. Safety and Feasibility of an Immersive Virtual Reality Intervention Program for Teaching Police Interaction Skills to Adolescents and Adults with Autism. Autism Research. 2020;13: 1418–1424. doi:10.1002/aur.2352

18. Green D, Charman T, Pickles A, Chandler S, Loucas T, Simonoff E, et al. Impairment in movement skills of children with autistic spectrum disorders. Dev Med Child Neurol. 2009;51: 311–316. doi:10.1111/j.1469-8749.2008.03242.x

19. Surgent OJ, Walczak M, Zarzycki O, Ausderau K, Travers BG. IQ and Sensory Symptom Severity Best Predict Motor Ability in Children With and Without Autism Spectrum Disorder. J Autism Dev Disord. 2021;51: 243–254. doi:10.1007/s10803-020-04536-x

20. Conners CK. Conners third edition (Conners 3). Los Angeles, CA: Western Psychological Services. 2008.

21. Jaime M, Longard J, Moore C. Developmental changes in the visual-proprioceptive integration threshold of children. J Exp Child Psychol. 2014;125: 1–12. doi:10.1016/j.jecp.2013.11.004

