## Supplement for "HaptiKart: An engaging videogame reveals elevated proprioceptive bias in individuals with autism spectrum disorder"

**Supplemental Document**

***Full Experimental Paradigm***

This study examines sensory-motor bias, derived from two counterbalanced conditions where visual and proprioceptive feedback were selectively perturbed (Bias Score = Error*_V+DP_* – Error*_DV+P_*). Since the assessment of sensory-motor bias assessment was the focus (and well controlled via counterbalancing), the full paradigm is detailed here in the supplement rather than the manuscript.

Each participant completed a ~20-minute paradigm consisting of perturbation conditions (6 laps each) followed by shorter washout blocks (3 laps). The washout blocks, which remained consistent throughout, provided veridical visual and proprioceptive feedback. The condition order was fixed for all participants, except for the counterbalanced conditions used to calculate bias scores.

Participants began with a *familiarization* block on a unique track (6 laps on track 1), where they experienced veridical visual and proprioceptive feedback to get accustomed to the steering wheel, videogame, and force feedback. Following familiarization, they completed six distinct perturbation conditions (6 laps on track 3), each separated by a washout block (3 laps on track 2). The perturbation conditions included:

1. Veridical visual without proprioceptive feedback (V)
2. Veridical visual with veridical proprioceptive feedback (V+P)
3. Rotated visual with veridical proprioceptive feedback (RV+P)
4. Delayed visual with veridical proprioceptive feedback (DV+P)
5. Veridical visual with delayed proprioceptive feedback (V+DP)
6. Delayed visual with delayed proprioceptive feedback (DV+DP)

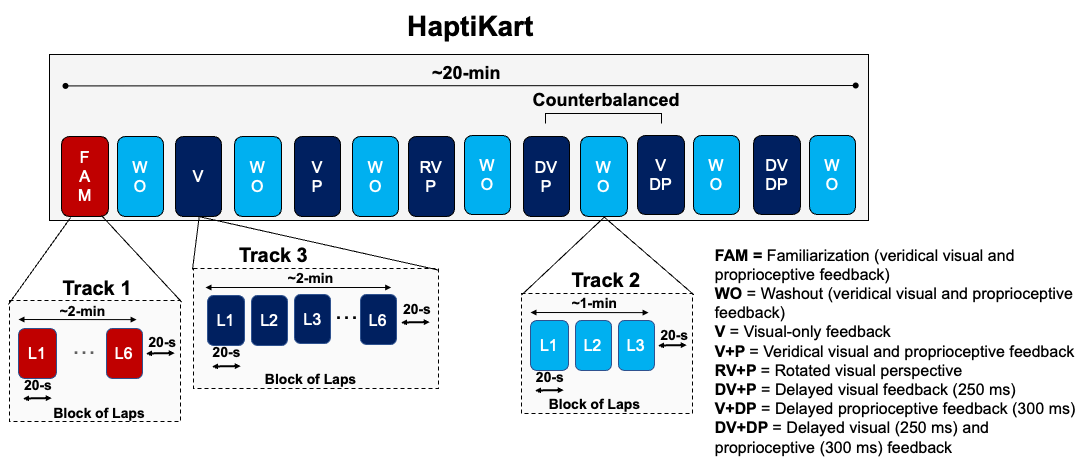

**Figure S1**: Experimental paradigm of the full HaptiKart paradigm. Different colors indicate the track used in the block (Track 1 [red] = familiarization block; Track 2 [light blue] = washout blocks; Track 3 [dark blue] = perturbation blocks).

***Lap Times***

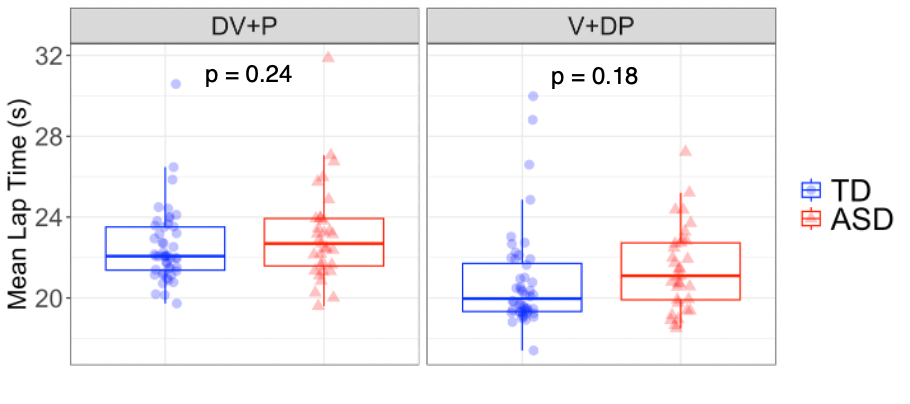

**Figure S2:** Lap times (seconds) for each participant averaged across six laps and displayed by group and condition.

***Steering Wheel Parameters***

The Logitech G29 steering wheel settings included a spring coefficient of 10%, spring saturation of 50%, and damper force of 10%, providing proprioceptive-error feedback that guided the participant while necessitating steering input to remain on the track centerline. This is demonstrated below in **Figure S3**, which shows the driving errors produced by the car without user input (i.e., self-driving). The feedback we provided in this study created a balance of guidance that continued to require user input to follow the track centerline. In task-development piloting conducted prior to the study, we determined that a constant *k* value of 1.2, applied separately to both heading and position errors, generated an appropriate level of proprioceptive-error feedback, ensuring that force feedback was directly proportional to the participant’s deviation in steering or position. The stiffness of the steering wheel remained constant throughout the experiment (i.e., 10% spring coefficient), meaning that the force experienced by participants increased linearly with their positional and heading errors. This setup provided intuitive feedback to encourage corrective steering adjustments. These parameters were controlled using a configuration file and the Logitech SDK.

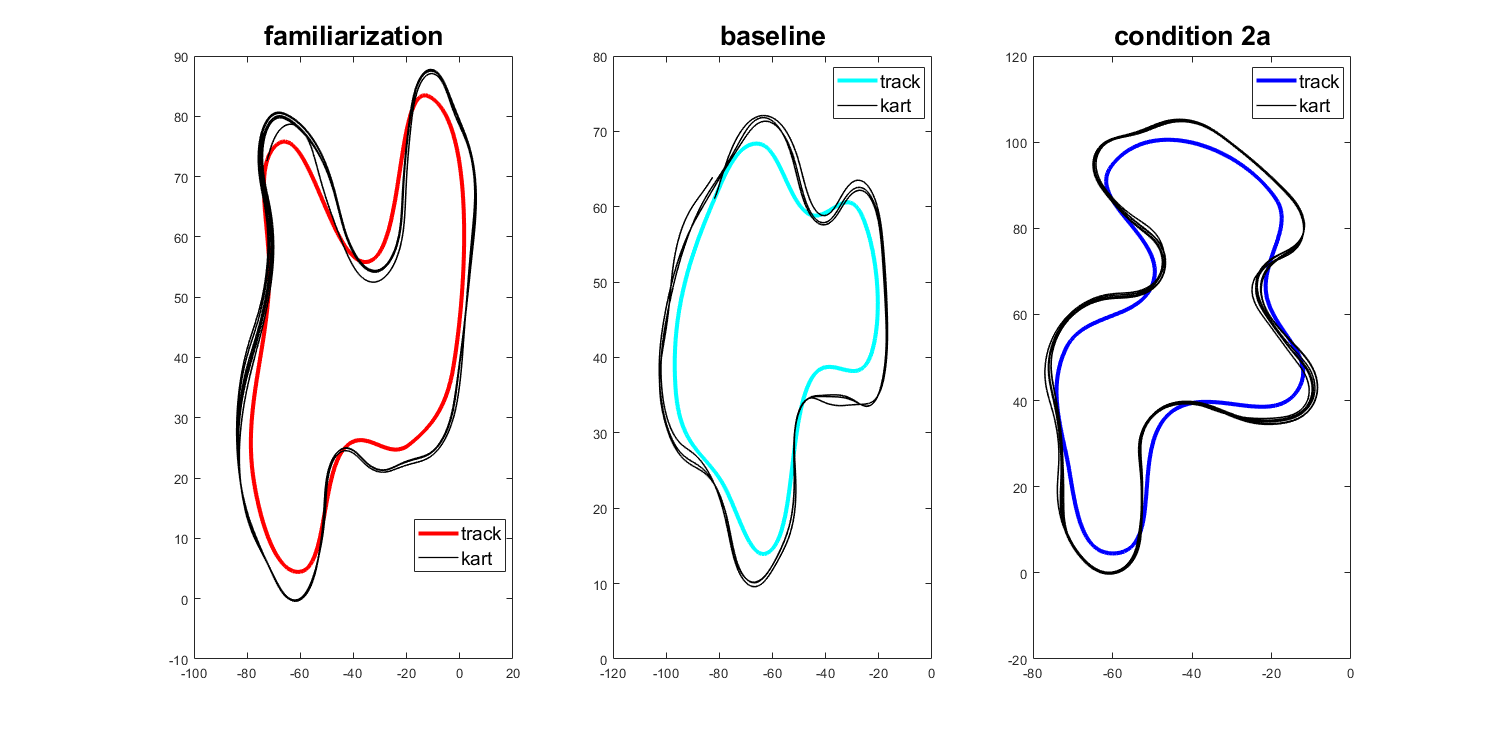

**Figure S3**: Driving path of car (black lines) over 6 laps produced by proprioceptive-error feedback without any user input (i.e., self-driving). The track centerline is shown in blue.

***Online Learning Effects***

Online learning effects were examined separately for visual delay (DV+P) and proprioceptive delay (V+DP) conditions. Independent samples t-tests revealed no significant between group differences in online learning for either delay condition (**Figure S4**).

$$Online Learning (\%)=\left( \frac{Initial Lap Error-Final Lap Error}{Initial Lap Error} \right)\times100$$

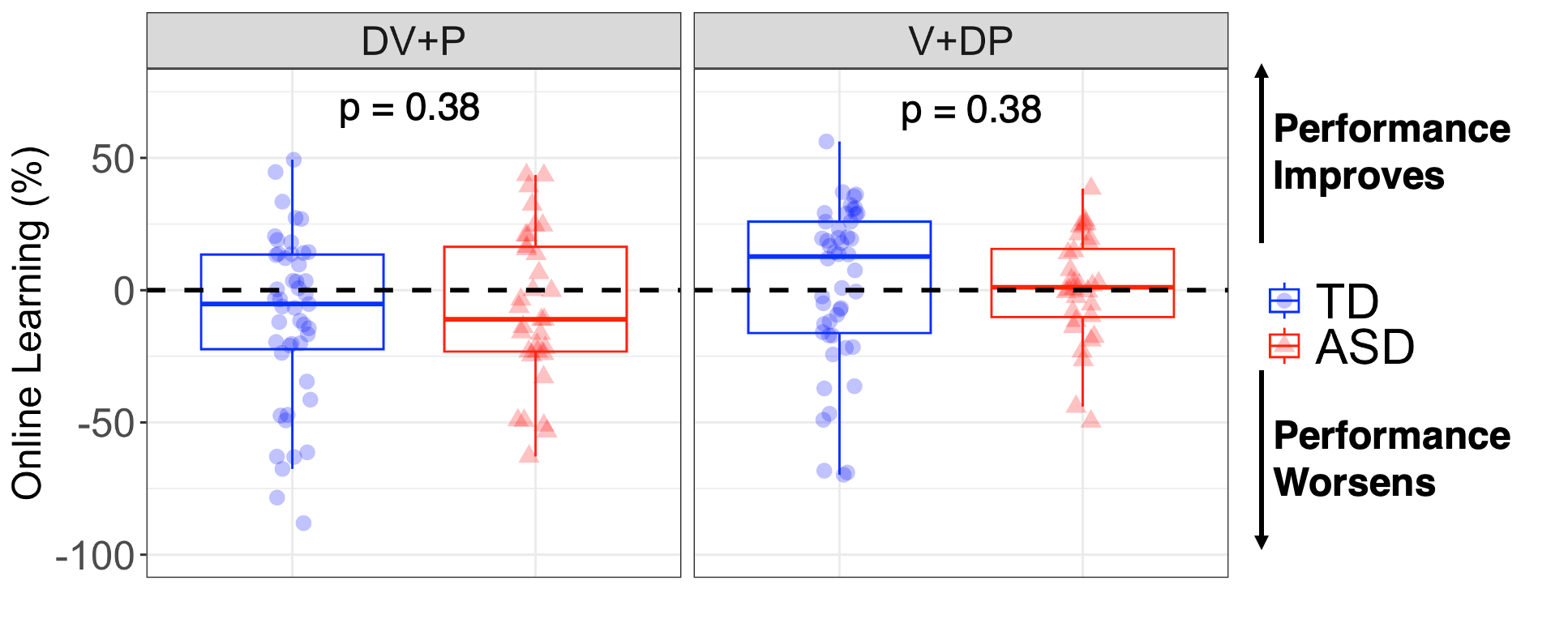

**Figure S4:** Online learning did not significantly differ between groups for visual or proprioceptive delay conditions (both *p* = 0.38).

**Track Generation**

*Bezier Curve Track Generation in Unity*

In HaptiKart, tracks were manually created using Bezier curves, as shown in **Figure S5**. Control points (red and blue dots) determine the shape of the curve, enabling smooth, continuous track paths. Multiple Bezier curve segments were connected to form the full track, allowing for precise control over the track design, which includes sharp turns and smooth sections. This method ensured that the tracks remained consistent and navigable. Three distinct tracks were developed using this approach and used in the full paradigm (**Figure S6**).

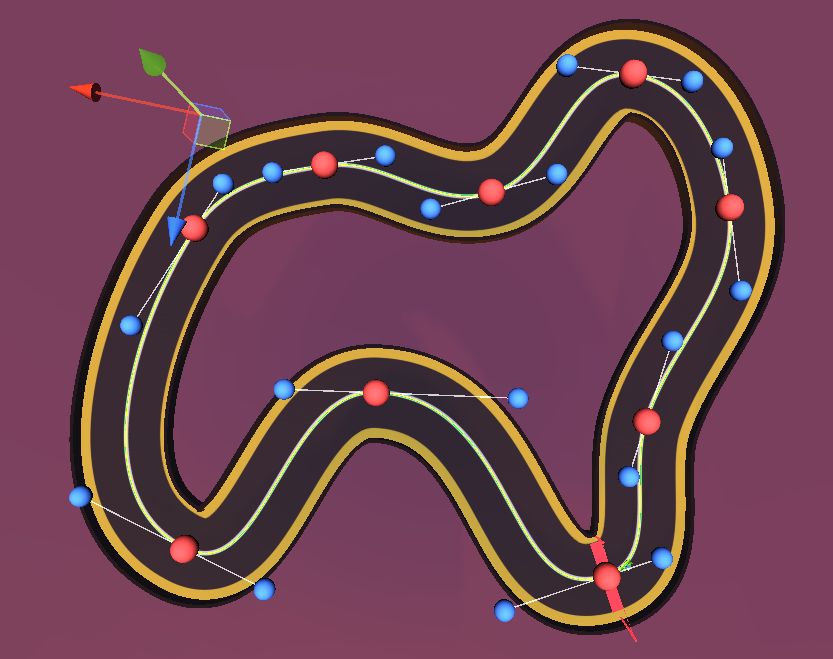

**Figure S5:** Bezier curve track generation in Unity

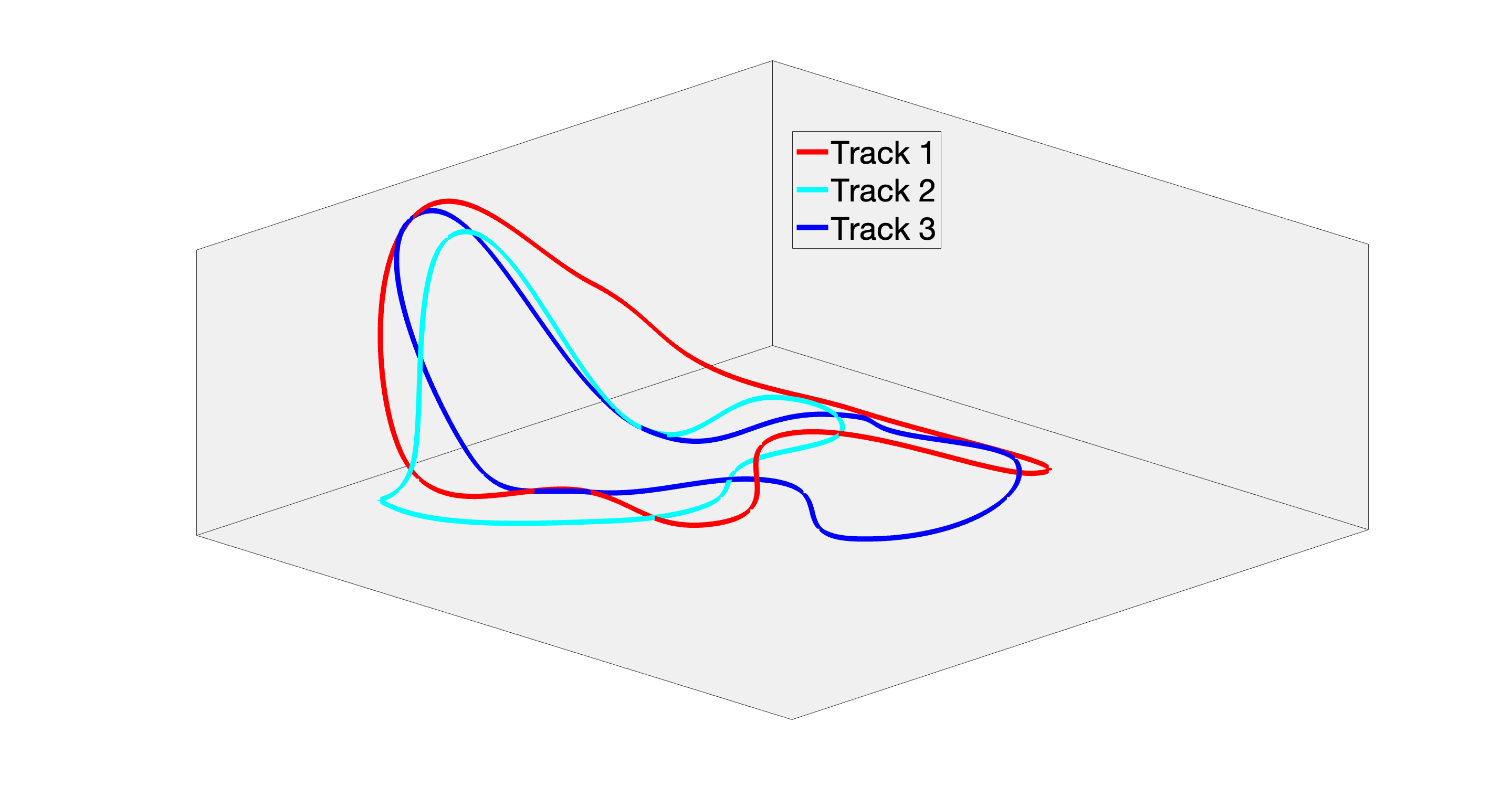

**Figure S6:** Track configurations used in the full paradigm.
